# A flexible formula for incorporating distributive concerns into cost-effectiveness analyses: priority weights

**DOI:** 10.1101/19003780

**Authors:** Ø.A. Haaland, F. Lindemark, K.A. Johansson

**Affiliations:** Bergen center for ethics and priority setting (BCEPS), Department of global public health and primary care, University of Bergen, Norway; Department of addiction medicine, Haukeland university hospital, Bergen, Norway; Department of international collaboration, Haukeland university hospital, Bergen, Norway

**Author notes:** Corresponding author: Øystein Ariansen Haaland, PhD, PO Box 7804, NO-5018 Bergen. Other authors: Frode Lindemark, Kjell Arne Johansson.

## Abstract

Cost effectiveness analyses (CEAs) are widely used to evaluate the opportunity cost of health care investments. However, few functions that take equity concerns into account are available for such CEA methods, and these concerns are therefore at risk of being disregarded. Among the functions that have been developed, most focus on the distribution of health gains, as opposed to the distribution of lifetime health. This is despite the fact that there are good reasons to give higher priority to individuals and groups with a low health adjusted life expectancy. Also, an even distribution of health gains may imply an uneven distribution of lifetime health.

We develop a systematic and explicit approach that allows for the inclusion of lifetime health concerns in CEAs, by creating a new priority weight function, PW=α+(t-γ)·C·e^-β·(t-γ)^, where t is the health measure. PW has several desirable properties. First, it is continuous and smooth, ensuring that people with similar health characteristics are treated alike. Second, it is flexible regarding shape and outcome measure, so that a broad range of values may be modelled.

Third, the coefficients have distinct roles. This allows for the easy manipulation of the PW’s shape. In order to demonstrate how PW may be applied, we use data from a previous study and estimated the coefficients of PW based on two approaches.

The first considers the mean weights assigned by the respondents:

PW_mean_=-0.42+(t+22.2)·0.27·e^-0.031·(t+22.2)^.

The second considers the median weights:

PW_median_=0.79+(t+8.85)·0.17·e^-0.053·(t+8.85)^.

This illustrates that our framework allows for the estimation of PWs based on empirical data.

## INTRODUCTION

Prioritisation of limited health care resources involves saying no and yes, hard ethical problems and reasonable disagreements. In health economics, cost-effectiveness analysis (CEA) is used to identify the most efficient allocation of health care resources. Within such a framework, informed decisions based on explicit CEA rankings can be made. However, CEAs have been extensively criticised for not being sensitive to fair distribution of health benefits, and there are other important equity concerns which may be considered, and few standardised methods are available to directly adjust incremental cost-effectiveness ratios (ICERs) [1-3]. CEAs are used to inform policy decisions on the introduction or reimbursement of new technologies or pharmaceuticals in countries such as the UK, Norway, Sweden, the Netherlands, New Zealand and Australia, in some cases including considerations beyond CEA through concepts like severity or burden of disease [4-14].

Typical health metrics in CEAs are quality-adjusted life years (QALYs) gained and disability-adjusted life years (DALYs) averted [15]. These aggregate measures take into consideration not only the life expectancy (LE) of a group, but also the quality of life. Interventions and health programs are typically ranked by ICERs, where those that maximize QALYs gained or DALYs averted are favoured. However, such rank-orders of interventions based on ICERs have been criticized because they disregard concerns for those who are worst off [16]. There are good reasons to give higher priority to individuals and groups with impaired health [16]. As opposed to health maximisation, one could aim for equal distribution of health across all individuals in society. This would require that resources should always be directed at people with the most impaired health, so as to minimize inequality in health outcomes, even if this means that the total health in the population will be reduced. People’s intuition about what is fair may lie somewhere between strict health maximisation and strict equality. A compromise between these perspectives is that health maximization is an important goal, but health gains to people with impaired health are assigned more weight than similar gains to people with better initial health [17]. This “ in between” position is called prioritarianism and has been defended by the philosophers like Derek Parfit [18] and Matthew Adler [19]. According to prioritarianism, a cheap and effective intervention directed at the healthiest may be preferred to an expensive and ineffective intervention directed at those who have worse health if left untreated. Still, if cost and effect were equal, the allocation of resources should be directed towards people with more impaired health if left untreated. Finally, when comparing competing health programs, studies have found that people favor priority to worse off, and they favor interventions that benefit those that are worse off over slightly less cost-effective interventions that benefit better off groups [20, 21]. Such equity concerns are too important to leave out from standard CEAs.

One may also argue that non-health concerns, like age, indirect benefits, productivity, and financial risk protection, are also important regarding equity. However, this paper focuses on health related concerns. At the time of a prospective health intervention, concerns for health distribution can be roughly divided into two perspectives: The first perspective is forward looking and is only concerned with the distribution of the health gains, typically measured in QALYs or DALYs. E.g., one may think that one QALY gained for 50 people is better, equal or worse than 50 QALYs gained for one individual. The second perspective is dealing with the distribution of health over a lifetime, typically measured as quality adjusted life expectancy from birth (QALE) [22, 23]. E.g., consider two individuals who may benefit from some treatment. One has QALE=80 without treatment and 81 with, and the other has QALE=20 without treatment and 21 with. Their gains would be equal, but the second is expected to get much less lifetime health. Hence, if an equal distribution of lifetime health in the population is a priority, one would value the set {80, 21} above {81, 20}. The view that expected lifetime health is what matters draws support from the literature on health priorities and some empirical studies [24-28]. Note that although QALE often increases with age, a high age does not necessarily correspond to a high QALE. E.g., the QALE of an old person who has experienced a lifetime of chronic illness may be far less than that of a healthy child. In this paper we focus on applications of the lifetime health perspective.

Our aim is the development of a systematic and explicit approach to empirically include these equity concerns in the framework of CEAs, by creating a new priority weight function. The function should be smooth and continuous in order to treat people with similar characteristics alike. Further, it should be flexible regarding shape and outcome measure to be able to encompass a broad range of values. Finally, coefficients should have distinct roles, to ensure that different functions can be easily compared, and allow for easy manipulation of the function’s shape.

## METHODS

### Moving from utility curves to priority weights

A social welfare function (SWF) is a function that represents the sacrifices that society is willing to make to promote a more equitable health distribution. The input should be a health distribution (individual QALYs, DALYs, etc.), and the output should be a real number, so that different distributions can be easily ranked. An SWF incorporates trade-offs between total health gain and health inequality in a population. Well-known SWFs are the Gini index and the Atkinson index [29, 30]. Both include a parameter that indicates the degree of inequality aversion, ranging from 0 to infinite. Such SWFs are data hungry, because complete data about the health or health gains for an entire population are needed. Often the data required does not even exist. A different set of SWFs is the one based on the aggregated QALY model. These utilitarian SWFs have the form

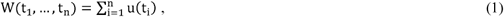

where t_i_ is the number of QALYs received by individual i (i=1,…,n), and the utility function u(t), defined over t_i_, is positive and monotonously increasing (u(t) ≥ 0, du/dt > 0). If u(t) is concave (d^2^u/dt^2^ < 0), W represents societal preferences for equality. Equality indifference yields u(t) = t, and reduces W to be the sum of the QALYs received across all individuals in society. Finally, if u(t) is convex (d^2^u/dt^2^ > 0), society is assumed to prefer inequality. The function u(t) has been the focus of several papers [29, 31-34]. As opposed to the Gini and Atkinson indexes, calculating u(t) does not require complete information about the health or health gains of an entire population (t_l_,…, t_n_). We refer to Rodriguez and Pinto for a more detailed discussion on u(t) [34].

However, u(t) does not account for the distribution of lifetime health. We approach this problem by considering priority weight functions (PWs) instead. These are functions that adjust the weight of health gains according to health related equity concerns, such as health attainment in a lifetime perspective. A general form of an SFW based on PWs is

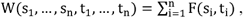

where t_i_ is the health level of individual i at the time of intervention, s_i_ is the health gain, and

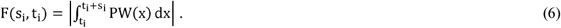

The vertical bars denote that the absolute value is to be considered, and x is a dummy variable used for integration. As an example of how (6) may be calculated, consider a sick individual with a health level without intervention of QALE=60, and an intervention with a gain of 3 QALYs. The priority weighted gain of the intervention is now 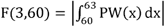.

Although the literature is lacking, one candidate for PW is the age-weight function of the Global burden of disease (GBD),

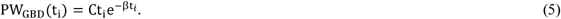

Here t_i_ is the age of individual i [35]. As mentioned, age weights are beyond the scope of this paper. Still, by letting t_i_ denote the health gain or lifetime health of individual i, (5) becomes a PW with quite an appealing form, although lacking in flexibility.

Flexibility is an important property of a PW, as it should be able to reflect a wide range of health related equity concerns. Further, if the coefficients of the PW have distinct roles, they can easily be adjusted to accommodate such concerns. E.g., one may wish to set an upper limit for the maximum weight, or set the maximum weight at a fixed value of t. Also, if the PW is fit to empirical data on people’s preferences, the roles of the coefficients will help to disentangle different concerns of society. One may, e.g., wish to estimate both the t for which the PW reaches its maximum, and what the maximum is. This is not straightforward in (5). A PW should also be twice differentiable with respect to t, so that it is continuous (no jumps) and smooth (no breaking points), ensuring the approximately equal treatment of individuals with similar values of t.

### A priority weight function

In this section we present a new flexible PW with coefficients that have distinct roles, which is suitable in a lifetime health framework via (6). The formula is as follows:

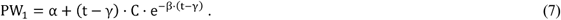

Setting α= γ= 0 reduces (7) to (5). Setting C = 0.1658 and β = 0.04 we get the weights shown in Figure 1A. Panels B and C in Figure 1 show how PW_1_ can be altered by manipulating C and β. As seen, C has the effect of inflating or deflating PW_1_ (Figure 1B). For β> 0, it can be shown that (7) ultimately approaches α with increasing t. We see that for larger β’s this happens more quickly than for smaller β’s (Figure 1C). When β ≤ 0, (7) increases with t, and never approaches α at all.

**Figure 1.**
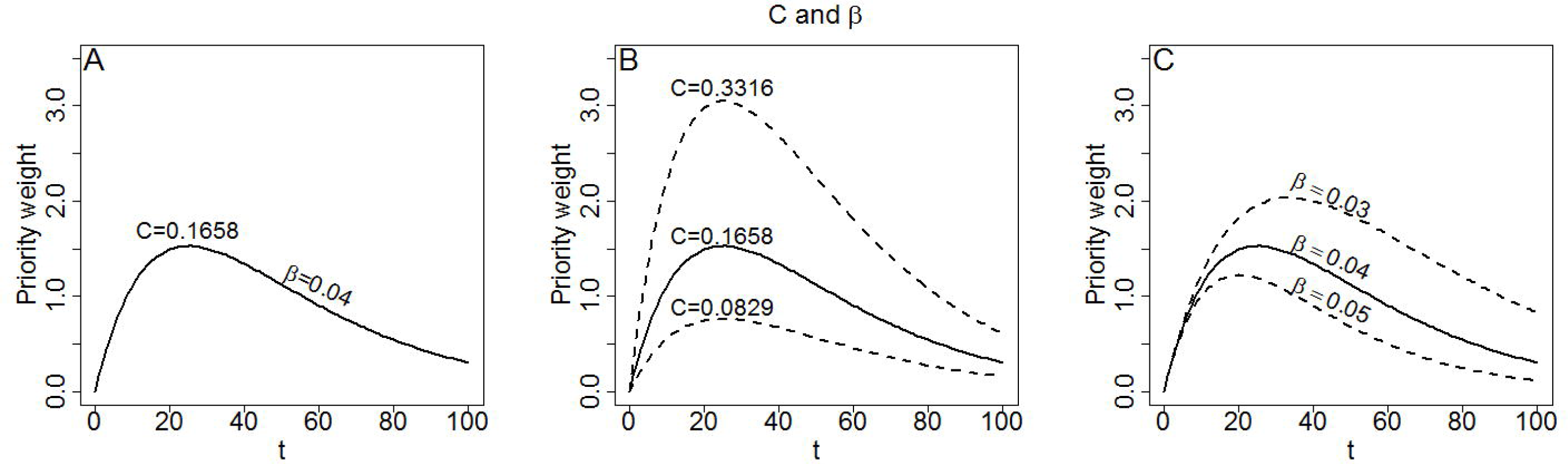
Properties of the coefficients of (7) when α= γ = 0. Panel A: Priority weights according to t. Panel B: Altering C causes an inflation or deflation. Panel C: Altering β changes the “pointyness” of the hump, causing the peak to become more or less pronounced.

The intercept α of (7) shifts the curve up or down (Figure 2A), whereas γ provides a right-left shift (Figure 2B). As is shown in Figure 2B, the maximum can be moved beyond t= 0, making (7) strictly decreasing. Note that C is interpreted slightly differently for (7) than for (5). For (7), changing C will affect the relative weights between different values of t unless α= 0. Also, C ≤ 0 is not a problem for (7), because α can be adjusted to make sure that PW_l_ >0 for all relevant t’s. A negative C simply causes (7) to be reflected through the line PW_l_ = α, or in other words, turned up-side-down. If C= 0, (7) reduces to the line PW_l_ = α, corresponding to inequality indifference. This is also true if β approaches infinity.

**Figure 2.**
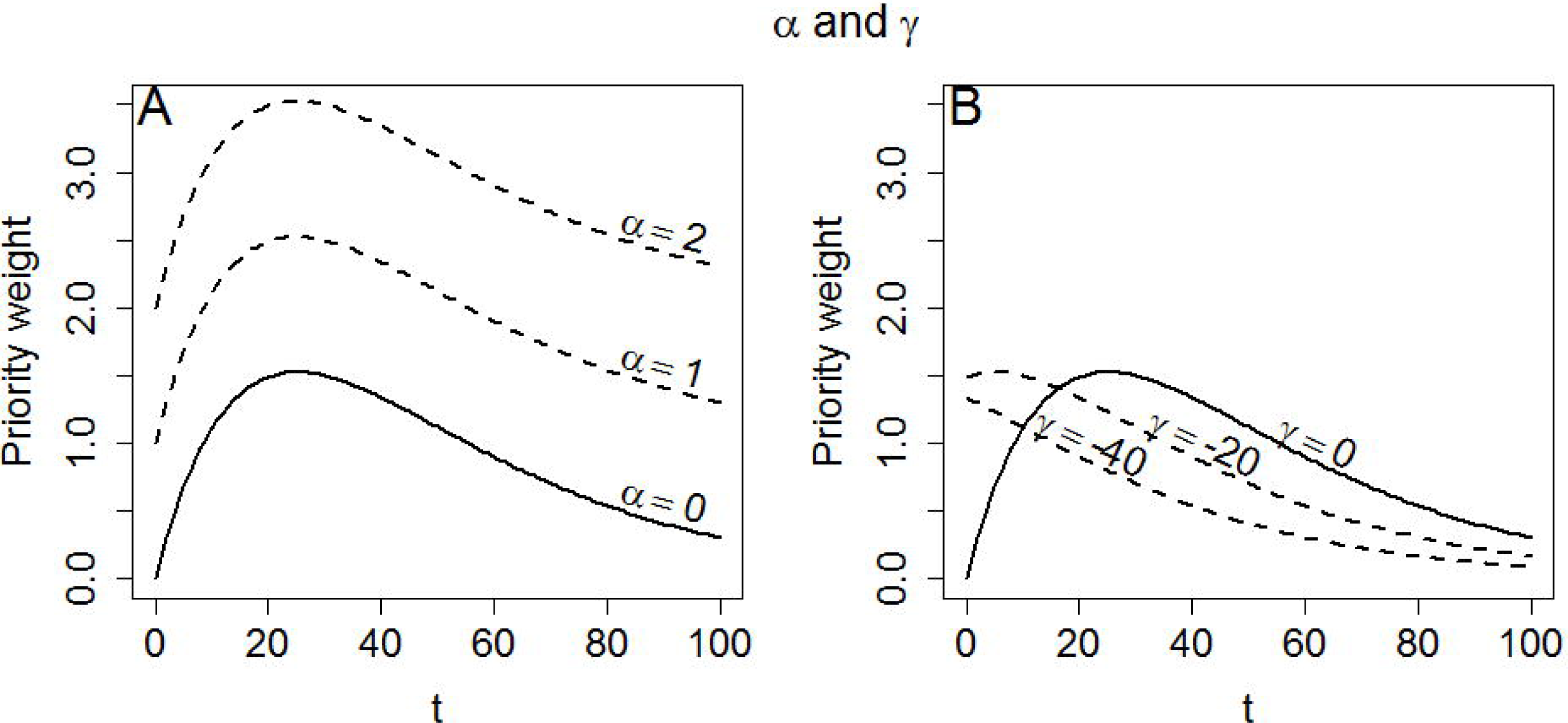
Properties of the coefficients of (7) when C = 0.1658 and β = 0.04. Panel A: Altering α causes an upward or downward shift. Panel B: Altering γ causes a left or right shift.

As demonstrated in Figure 1 and Figure 2, manipulating the different coefficients of (7) allows us to flexibly change its shape. E.g., if we want to ensure that people with a certain value of t are given a certain weight (anchoring the curve), we may simply change α. Letting

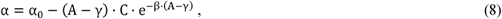

anchors the curve to α_0_ at t= A. Further, if we want to set a maximum weight, so that max (PW_l_)= w_m_, we let

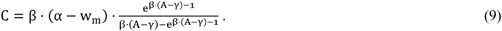

If α_0_, A, γ, w_m_ and β are given, we can treat (8) and (9) as two linear equations with two unknowns, and solve for C and α. Figure 3A shows (7) anchored to 1 at t= 80 for different values of w_m_.

**Figure 3.**
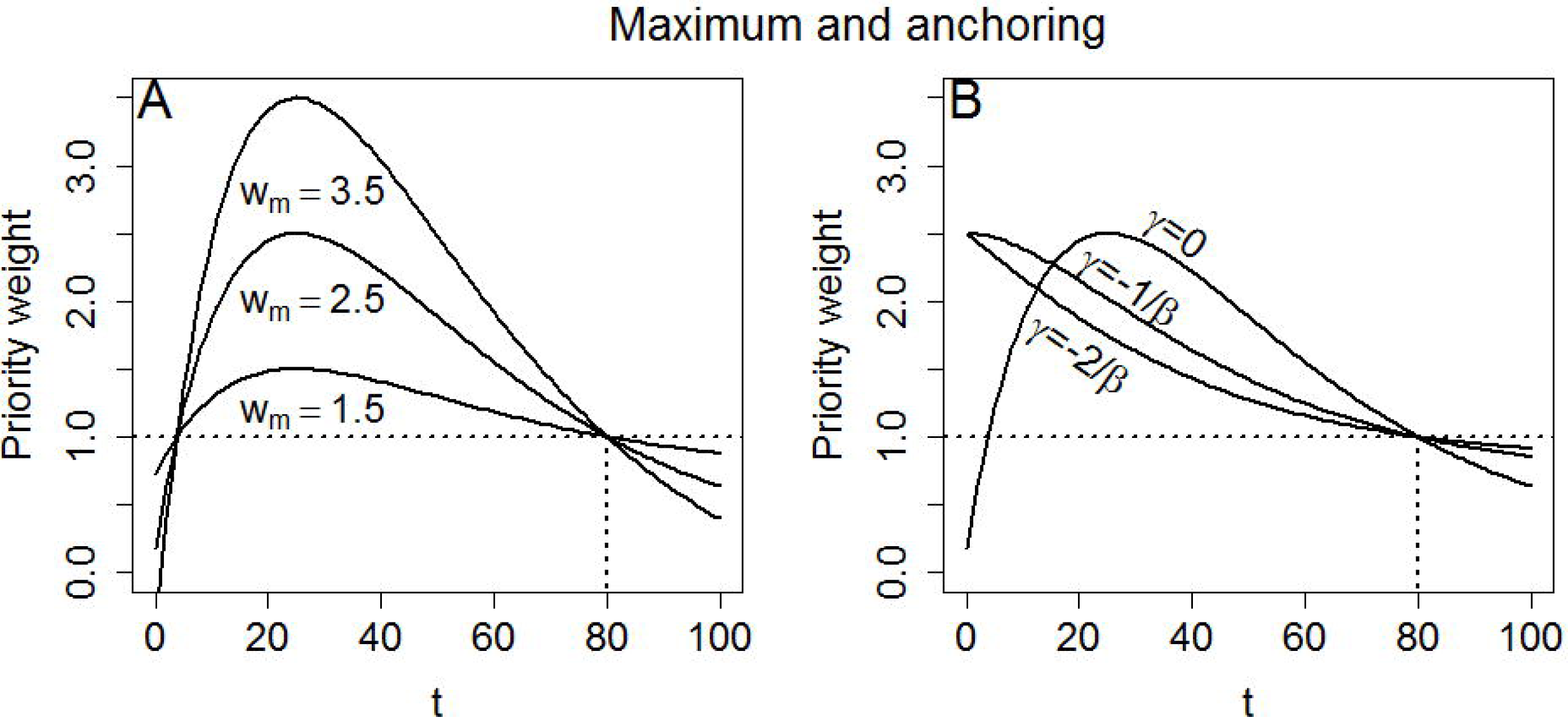
Anchoring to 1 for t= 80, with β = 0.04. Panel A: Varying max t (w_m_) when γ = 0. Panel B: Varying γ when w_m_ = 2. 5. For γ = o, there is no shift. For γ = -1/ β the curve is shifted left so that the maximum is at t= 0. For γ = -2/ β the curve is shifted so that the slope is the steepest at t= 0.

As mentioned, manipulating γ will shift (7) right or left. It may be of particular interest to identify the shift needed to ensure that the maximum is moved to t= 0. This means that the slope is flat at t= 0, and gradually becomes steeper, so that gains for most individuals with poor lifetime health are given weights close to w_m_. Differentiating (7) with respect to t, we find that the maximum is when t= 1/β. Hence, letting γ= -1/β ensures that the maximum is at t=0 (Figure 3B). Differentiating (7) once more with respect to t, the t for which the slope is the steepest can be identified as 2/β. Therefore, letting γ= -2/β causes (7) to be at its steepest when t= 0. In such a scenario, only gains to people with very poor health are given weights close to w_m_ (Figure 3B).

So far the mathematical properties of the coefficients in (7) have been presented, but the policy implications and conceptual meaning of these properties may not be obvious. The next paragraphs are dedicated to the discussion of these implications. First, we consider γ. If γ> -1/ β, it is possible that PW(t_l_)< PW(t_2_) for t_l_ < t_2_. This makes sense if one thinks that small gains to individuals with almost no lifetime health should be worth less than a similar gain to an individual with more lifetime health. This could pertain to treatment of children born with very severe conditions. In Figure 1 and Figure 2 the default value of γ was 0.04, so that the maximum was at t = 1/0.04 = 25 QALYs, which is probably too high in most scenarios. When γ ≤ -1/ β, a gain of, e.g., 1 QALY will always be weighted higher for the individual with the least lifetime health. From now on we shall assume that γ ≤ -1/ β.

Next we consider α. The particular value of α is not necessarily easy to interpret, but because this coefficient allows for the anchoring of the PW at a certain weight for a certain value of t, it is useful when selecting a reference value of t for which PW=1 (e.g., one may anchor PW at 1 when QALE=70). Once a reference is chosen, the interpretation of the other coefficients is easier. A large β corresponds to a narrow peak of the PW, ensuring a steep decline of the curve. Now, gains to individuals with little lifetime health are given much higher weights than gains to individuals with more lifetime health, whereas gains to individuals with intermediate and high levels of lifetime health will be weighted similarly. A small β yields a flat peak, so that the weight drops steadily as the initial health level improves (t increases). Although β also will affect the range of possible weights, this property is more closely connected to the C. The choice of C decides how much value will be assigned to a given t relative to the reference. E.g., one may think that the maximum weight should be no more than three times the weight of the reference. A very small C would ensure that all health gains were treated almost the same.

### Application of the priority weight function to experimental data

In order to illustrate the potential use of (7) in a CEA within a lifetime health framework, we fit the function to data from a discrete choice experiment. The procedure involves two conceptually simple steps. First, data must be collected, and then the curve must be fit. However, both these steps may be performed a number of different ways. We shall focus on an approach that is suitable for a dataset presented in Ottersen et al. (2014).

Ottersen et al. used a computer based questionnaire to ask 96 students in Norway how they valued QALYs gained for people at different initial health levels, measured in “ initial QALE” (QALE without intervention). The reference case was a group of healthy people with QALE=70 who were subject to an intervention that would give them 10 extra QALYs (i.e., move from QALE=70 to QALE=80). Respondents were asked to assign QALYs to individuals with worse initial health (QALEs in the set {10, 25, 40, 55}) in such a way that they were indifferent between the different scenarios. Now, priority weights could be calculated for each individual respondent based on the ratio between the QALYs assigned to the unhealthy groups and the reference group. E.g., consider a respondent who was indifferent between a situation where the group with initial QALE=25 gained 5 QALYs (i.e., moving from 25 to 30) and the reference group gained 10 QALYs (i.e., moving from 70 to 80). This would mean that gains in the group with a worse initial health, where initial QALE=25, are regarded as more important than gains in the healthy group, where initial QALE=70. Mathematically, this extra weight can be expressed as V(25)=10/5=2 relative to the reference group (QALE=70). We refer to Ottersen et al. for a more thorough discussion of data and study design.

One may organize the data as follows. Two lists, V and T, are constructed. V contains all the weights assigned by the respondents, and T contains the initial QALEs, so that the weight V_i_ corresponds to the QALE T_i_. For purposes of estimating the coefficients of (7), we treat the observations as independent. Applying a least squares approach, we get that the sum of squared errors (SSE) is

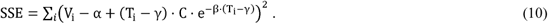

This expression may be minimized using a numerical routine or by solving the set of equations

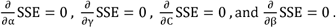

If one wishes to anchor the curve to a specific value (e.g., the data from Ottersen et al. has V(70)=1), it is straightforward to use (8) in (10). Setting maximum weights would imply using (9) in (10). Also, any coefficient in (10) may be kept constant.

Minimizing SSE ensures that the curve is fit to the mean V for each T in {10, 25, 40, 55}. Using the R function `optim()` to minimize SSE, we get

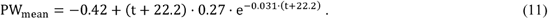

However, minimizing instead the sum of absolute errors,

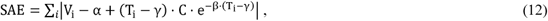

the curve is fit to the median. Now PW becomes

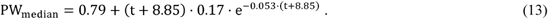

Note that when fitting the curves, α and γ were set to ensure that PW(10) was the maximum (i.e., PW(10)=w_m_) and PW(70)=1. Figure 4 shows (11) and (13) plotted with the empirical data. Note that the PW_mean_ is more affected by extreme values than are PW_median_.

**Figure 4.**
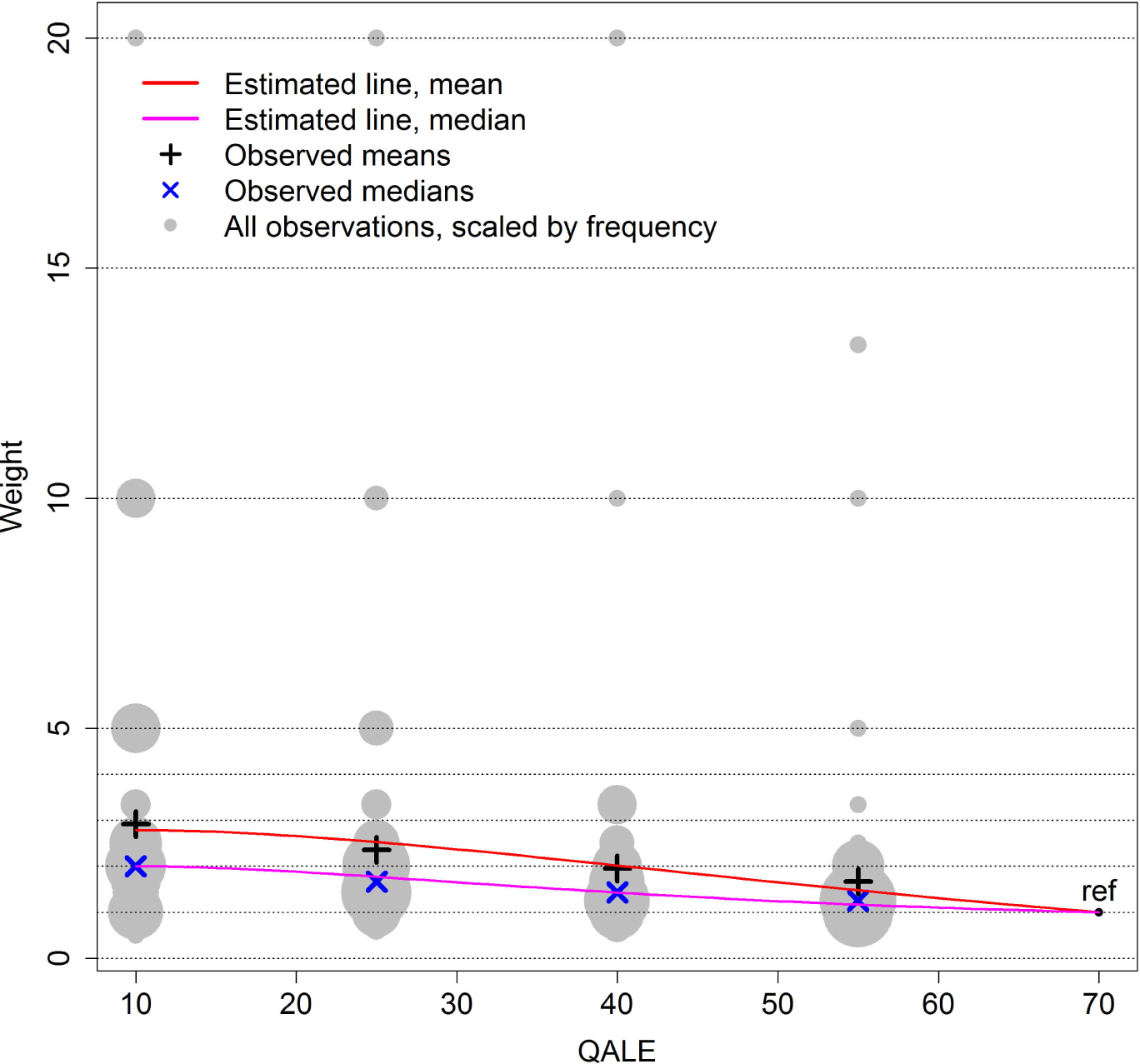
Estimating PW from data. The red line minimizes (10) to estimate a PW based on the means (black +). The coefficients are *α* = -0.42, *γ* = -22.2, *C* = 0.27, and *β* = 0.031. The pink line minimizes (12) to estimate a PW based on the medians (blue x). The coefficients are *α* = 0.79, *γ* = -8.85, *C* = 0.17, and *β* = 0.053. In both scenarios PW(10)=w_m_ (maximum weight at T=10) and PW is anchored at PW(70)=1.

### Data availability statement

All necessary data and code to reproduce Figure 4 are available at https://github.com/oeh041/Priority_weights and https://doi.org/10.5281/zenodo.3256151

## DISCUSSION

We have presented a new priority weight function (PW) with key characteristics that are important in consistent priority setting across patient groups and populations. As demonstrated, the PW was suitable for CEA in a lifetime health framework. The PW is flexible and easy to adjust to a broad range of equity considerations by altering a limited number of comprehensible parameters, each with its distinct role. We may set the range of priority weights by modifying C, we may shift the curve upwards or downwards by modifying α, we may shift the curve left or right by modifying γ and we may set the “ pointiness” of the curve by modifying β. These characteristics make it easy, e.g., to anchor the curve or to set maximum weights. Considering (11) and (13) (Figure 4), we see that β_median_ > β_mean_, indicating that PW_median_ decreases at a faster rate than PW_mean_ (i.e., PW_median_ is pointier). Further, we have that C_median_ < C_mean_, meaning that PW_mean_ adds more weight to the worse off than does PW_median_.

Drawing the PW by hand would of course be more flexible than using (7). However, we argue that setting some restrictions for the behaviour of the PW is useful. First, it offers consistency when comparing different curves. Second, the space of possible PWs is limited by (7), but the major restrictions caused by (7) seem reasonable (smooth and maximum one hump). Hence, a large number of nonsensical PWs are impossible (discontinuous and multiple humps).

We did not account for discounting of future health. It is not clear how this would affect rankings of interventions. A positive discount rate would put different weights on equally sized gains that were experienced at different times from the start of intervention (e.g., 0.5 QALYs gained immediately is valued higher than 0.1 QALYs gained each year in five years, or 0.5 QALYs five years from now). Hence, interventions with immediate effects would be valued over interventions where the benefit occurs in the future. Also, discounting health gains would give a relatively low priority to those with large gains, who are expected to have less lifetime health. This is counterintuitive in a lifetime health priority perspective.

Although we do give an example of how priority preferences can be inferred from data, future work includes developing methodology to do inference about parameter estimates based on data from discrete-choice experiments. This would allow for the testing of differences in preferences between groups. E.g., conditioning on the other variables, one could test if men and women preferred different values of C (determining the maximum weight) or γ (determining the t for which the maximum weight was obtained). When an appropriate PW (or a range of PWs) has been selected, it could be applied to evaluate the equity impact of different interventions on reduction of mortality or morbidity. Also, work should be done to include non-health concerns like socioeconomic status and productivity in the model, and to explore the consequences of priority-weighted CEAs across settings.

In this paper we do not intend to determine which PW is the most appropriate, but rather to illustrate what can be achieved by a flexible general PW. However, we would like to stress the fact that a flat PW is also a PW. In other words, there is no such thing as “ not using a PW” in CEA. As illustrated, our framework allows for the estimation of PWs based on empirical data.

## Data Availability

All necessary data and code are available at https://github.com/oeh041/Priority_weights and https://doi.org/10.5281/zenodo.3256151

https://doi.org/10.5281/zenodo.3256151

## ACKNOWLEDGMENTS

We would like to acknowledge Ole Frithjof Norheim and Trygve Ottersen for their useful comments during the creation of this manuscript.

## REFERENCES

1. Johri M, Norheim OF: Can cost-effectiveness analysis integrate concerns for equity? Systematic review. Int J Technol Assess Health Care 2012, 28(2):125–132.

2. A QALY is a QALY is a QALY - or is it not? In: Measuring and valuing health benefits for economic evaluation. Edited by Brazier J, Ratcliffe J, Salomon JA, Tsuchiya A. Oxford: Oxford University Press; 2007: 287–298.

3. Baltussen R, Youngkong S, Paolucci F, Niessen L: Multi-criteria decision analysis to prioritize health interventions: Capitalizing on first experiences. Health Policy 2010, 96(3):262–264.

4. National Institute for Health and Clinical Excellence: Guide to the methods of technology appraisal 2013. In. London; 2013.

5. Claxton K, Sculpher M, Palmer S, Culyer AJ: Causes for concern: is NICE failing to uphold its responsibilities to all NHS patients? Health Econ 2015, 24(1):1–7.

6. Cookson R: Justice and the NICE approach. J Med Ethics 2015, 41(1):99–102.

7. Morland B, Ringard A, Rottingen JA: Supporting tough decisions in Norway: a healthcare system approach. Int J Technol Assess Health Care 2010, 26(4):398–404.

8. Svensson M, Nilsson FO, Arnberg K: Reimbursement Decisions for Pharmaceuticals in Sweden: The Impact of Disease Severity and Cost Effectiveness. Pharmacoeconomics 2015, 33(11):1229–1236.

9. van de Wetering EJ, Stolk EA, van Exel NJ, Brouwer WB: Balancing equity and efficiency in the Dutch basic benefits package using the principle of proportional shortfall. Eur J Health Econ 2013, 14(1):107–115.

10. Franken M, Stolk E, Scharringhausen T, de Boer A, Koopmanschap M: A comparative study of the role of disease severity in drug reimbursement decision making in four European countries. Health Policy 2015, 119(2):195–202.

11. Grocott R, Metcalfe S, Alexander P, Werner R: Assessing the value for money of pharmaceuticals in New Zealand--PHARMAC’s approach to cost-utility analysis. N Z Med J 2013, 126(1378):60–73.

12. Ngo P: The Influence of Cost-Effectiveness Evaluations on Reimbursement in Australia: A Retrospective Study of Decisions made by the Pharmaceutical Benefits Advisory Committee. Pharmaceutical Medicine 2014, 28(4):187–193.

13. Norges offentlige utredninger. Åpent og rettferdig - prioriteringer i helsetjenesten. NOU 2014:12. In: Norges offentlige utredninger; NOU 2014:12. 218 s.

14. Paulden M: Recent amendments to NICE’s value-based assessment of health technologies: implicitly inequitable? Expert Rev Pharmacoecon Outcomes Res 2017, 17(3):239–242.

15. Drummond MF, Sculpher M, Torrance G, O’Brien B, Stoddart G: Methods for the economic evaluation of health care programmes, 3rd ed. edn. Oxford: Oxford University Press; 2005.

16. Brock DW, Wikler D: Ethical Issues in Resource Allocation, Research, and New Product Development. In: Disease Control Priorities in Developing Countries. edited by Jamison DT, Breman JG, Measham AR, Alleyne G, Claeson M, Evans DB, Jha P, Mills A, Musgrove P, 2nd edn. Washington (DC): World Bank; 2006:259–270.

17. Cookson R, Drummond M, Weatherly H: Explicit incorporation of equity considerations into economic evaluation of public health interventions. Health Econ Policy Law 2009, 4(Pt 2):231–245.

18. Parfit D: Equality and priority. Ratio 1997, 10(3):202–221.

19. Adler MD, Holtug N: Prioritarianism: A response to critics. Polit Philos Econ 2019, 18(2):101–144.

20. Choudhry N, Slaughter P, Sykora K, Naylor CD: Distributional dilemmas in health policy: large benefits for a few or smaller benefits for many? Journal of health services research & policy 1997, 2(4):212–216.

21. Olsen JA: A note on eliciting distributive preferences for health. J Health Econ 2000, 19(4):541–550.

22. Ottersen T: Lifetime QALY prioritarianism in priority setting. J Med Ethics 2012.

23. Lindemark F, Norheim OF, Johansson KA: Making use of equity sensitive QALYs: a case study on identifying the worse off across diseases. Cost effectiveness and resource allocation : C/E 2014, 12:16.

24. Williams A: Intergenerational equity: an exploration of the ‘fair innings’ argument. Health Econ 1997, 6(2):117–132.

25. Norheim OF, Asada Y: The ideal of equal health revisited: definitions and measures of inequity in health should be better integrated with theories of distributive justice. Int J Equity Health 2009, 8:40.

26. Dolan P, Tsuchiya A: It is the lifetime that matters: public preferences over maximising health and reducing inequalities in health. J Med Ethics 2012, 38(9):571–573.

27. Bleichrodt H, Doctor J, Stolk E: A nonparametric elicitation of the equity-efficiency trade-off in cost-utility analysis. J Health Econ 2005, 24(4):655–678.

28. Ottersen T, Maestad O, Norheim OF: Lifetime QALY prioritarianism in priority setting: quantification of the inherent trade-off. Cost Eff Resour Alloc 2014, 12(1):2.

29. Atkinson AB: On the measurement of inequality. J Econ Theory 1970, 2(3):244–263.

30. Bleichrodt H, van Doorslaer E: A welfare economics foundation for health inequality measurement. J Health Econ 2006, 25(5):945–957.

31. Wagstaff A: Health care: QALYs and the equity-efficiency tradeoff. In: Cost-benefit analysis. Edited by Layard PRG, Glaister S, 2nd edn. Cambridge: Cambridge University Press; 1994 428–447.

32. Bleichrodt H: Health utility indices and equity considerations. J Health Econ 1997, 16(1):65–91.

33. Olsen JA: Persons vs years: two ways of eliciting implicit weights. Health Econ 1994, 3(1):39–46.

34. Rodriguez-Miguez E, Pinto-Prades JL: Measuring the social importance of concentration or dispersion of individual health benefits. Health Econ 2002, 11(1):43–53.

35. Murray CJ: Rethinking DALYs. In: The global burden of disease: a comprehensive assessment of mortality and disability from diseases, injuries, and risk factors in 1990 and projected to 2020. edited by Murray CJL, Lopez AD. Cambridge, MA: Published by the Harvard School of Public Health on behalf of the World Health Organization and the World Bank; Distributed by Harvard University Press; 1996: 1–98.

